# The prevalence and associated factors of adverse pregnancy outcomes among Afghan women in Iran; findings from community-based survey

**DOI:** 10.1101/2020.08.05.20168708

**Authors:** Omid Dadras, Takeo Nakayam, Masahiro Kihara, Masako Ono-Kihara, Seyedahmad Seyedalinaghi, Fateme Dadras

## Abstract

**Backgrounds:** An estimated 2.5 million Afghans are living in the Iran and almost half of them are young women at the childbearing ages. Although the evidence indicates lower rates of antenatal care and higher incidence of pregnancy complications in Afghan compared to Iranian women, the underlying reasons are not well defined. Therefore, in the present study, we aimed to explore the prevalence and associated sociodemographic factors of adverse pregnancy outcomes and examine the impact of intimate partner violence, food insecurity, poor mental health, and housing issues on pregnancy outcome in Afghan women living in Iran.

**Methods:** In July 2019, we enrolled 424 Afghan women aged 18-44 years old using the time-location sampling at three community health centers in the south region of Tehran province. The data was collected through face to face interviews using the researcher-developed questionnaire. Using bivariate and multivariate analysis, the impact of poor antenatal care, intimate partner violence, food insecurity, and poor mental health was assessed on the incidence of adverse pregnancy outcome.

**Results:** More than half (56.6%) of Afghan women reported at least one pregnancy complication in their recent pregnancy. The results showed that undocumented, illiterate, and unemployed Afghan women with lower socioeconomic status are more likely to experience adverse pregnancy outcomes. Furthermore, we observed lower prevalence of adverse pregnancy outcomes among documented immigrants with health insurance compared to those with no health insurance. It is also been found that the food insecurity [Adjusted OR = 3.35, 95% CI (1.34-8.36)], poor antenatal care [Adjusted OR = 10.50, 95% CI (5.40-20.39)], intimate partner violence [Adjusted OR = 2.72, 95% CI (1.10-6.77)], and poor mental health [Adjusted OR = 4.77, 95% CI (2.54-8.94)] could adversely impact the pregnancy outcome and we observed higher incidence of adverse outcomes among those suffering from these situations.

**Conclusion:** To our knowledge, this is the first study that explored the prevalence and associated factors of adverse pregnancy outcomes and the impact of intimate partner violence, food insecurity, poor mental health on pregnancy outcome among Afghan women in Iran. Enhancing the psychosocial support and empowering Afghan women through expanding the social network and safety net should be a priority for the central government and international parties. Psychological counseling should be incorporated into routine maternity care for Afghan refugees. Access to free antenatal care is a right for all Afghan women and it should be facilitated by universal health insurance for all Afghans regardless of their legal status.

## Introduction

Afghans are the second-largest refugee population after Syrians worldwide (1). During the last decades, Iran has been one of the countries that received a large number of Afghan immigrants and refugees in the Middle East (2). Afghans account for 96% of registered refugees in Iran, comprising 3% (approximately 2.5 million) of the total Iran population (3). Less than half of this population are women; however, the majority of them are young, and at the childbearing ages (4). Refugees are one of the most vulnerable population in the host country. The feeling of abandonment and isolation in the host society could endanger their physical and mental health. They may also experience multiple discriminations, violence, or exploitation which may impair their mental and physical health (3). In Iran, Afghan population include immigrants and refugees who either have valid documents or enter the country illegally from east border; therefore, instead of using refugee or immigrant, we used documented or undocumented immigrants in this paper.

In Iran, Afghan women are commonly subjected to several hostile encounters such as intimate partner violence, limited access to maternity care, and financial and housing struggles (5, 6). These issues are more likely to be seen among those of younger age and lower education or socioeconomic status (7) and could adversely affect the health of mother and offspring in women of fertile ages during pregnancy (8). A growing body of evidence documented lower access to maternity care and consequently higher occurrence of adverse pregnancy outcomes among immigrant and refugee women in different countries (8-10). Financial constraints, inadequate health literacy, costly care, and lack of health insurance are some of the obstacles in receiving effective and adequate maternity care among Afghan women in Iran (6, 8, 11). Despite the abundance of evidence indicating the low access of Afghan women to maternity care in Iran, the impact of such low access on the pregnancy outcome is still unclear.

Several notable factors have been associated with poor reproductive health outcomes among Afghan women in Iran. Intimate partner violence (IPV) has been linked to poor reproductive health outcomes such as unwanted pregnancy and lower uptake of modern contraceptive methods among Afghan women; however, the possible impact on pregnancy outcome was not characterized in previous studies (5). A study in Iran indicated high prevalence of IPV among Afghans (79.8%). It is almost five times higher than the reported number in Iranian women (14.1%) (12). Food insecurity is also a very important determinant of health outcomes in an expectant mother (13, 14). Moderate to severe food insecurity has been documented in approximately 60% of Afghan households in Iran (15), this could lead to adverse pregnancy outcomes in Afghan women suffering from lack of adequate nutrients and vitamins and may harm the health of the new borne (13). The poorer mental health has also been linked to the higher incidence of adverse pregnancy outcomes such as low birth weight or preterm birth in several studies (16, 17); however, the evidence is missing in specific ethnic groups such as Afghans. It is important to define the impact of IPV, food insecurity, and poor mental health on pregnancy outcome, especially among vulnerable populations such as Afghan refugee women, to prevent adverse pregnancy outcomes.

Although the current evidence indicates a higher prevalence of adverse pregnancy outcomes among Afghan women compared to Iranian women (6, 8), there is a gap in knowledge regarding the determinants and associated factors of such outcomes among Afghan women living in Iran. Therefore, in the present study, we explored the associated sociodemographic factors and the potential impact of intimate partner violence, poor mental health, food insecurity, and housing issues on pregnancy outcomes among Afghan women in Iran.

## Methods

### Study setting

We carried out a cross-sectional study among Afghan women living in Iran from June 2019 to August 2019. The study setting was decided by consensus between experts at Tehran University of Medical Sciences and Iran Ministry of Health. The south region of Tehran where the highest number of Afghan nationals are currently living was selected as the study setting. This region, so-called Share-Rey is located in the south-east part of Tehran province. There are 15 urban community health centers in this region in which primary health care including antenatal care is provided.

### Sample size

Based on the literature review (16), with the conservative assumption of dependent variable prevalence not to be less than 50%, the confidence level of 95%, a relative precision of 5% points on each side, and a response rate of 90%, the sample size was calculated 424. The response rate was 100% and we had no dropout or withdrawal during the study.

### Sampling method

Afghan population, particularly the undocumented ones, are really hard to reach in Iran. Therefore, applying the time-location sampling (TLS) method. TLS is a method for sampling the hard-to-reach population in places and at times where they congregate rather than where they live (17). Following the TLS sampling procedure, in the beginning, the common places and times that Afghan females visit to receive maternity care were identified. Through a primary field investigation, we realized that the urban community health centers are the most popular places for Afghan women to visit for maternity care and morning time is the usual time of visit. Therefore, we identified all the community health centers in Shahre-Rey (15 centers) and selected three centers with the highest number of Afghan visitors. We visited these centers in the morning time during the working days of four consecutive weeks in July 2019. A sample of 424 Afghan women, who met the eligibility criteria and consent to participate, were recruited proportionately to the size of the Afghan population visiting the corresponding community health center.

### Inclusion and exclusion criteria

The eligibility criteria were being an Afghan national, aged 18-49 years, given birth or being pregnant during the year before the interview, and lived in Iran at least a year and during the time of pregnancy. We excluded those with physical disabilities that affect their access to maternity care, those who visited or stayed in Afghanistan during their pregnancy, those who suffer from infertility, and those who did not consent to participate.

### Instrument and variables

The questionnaire (supplementary 1) of the present study was developed based on a comprehensive literature review and consulting the experts at Kyoto University and Tehran University of Medical Sciences. The questionnaire collected the data on participants’ sociodemographic characteristics including the age, education, employment, family income (<4million vs ≥4million-Iranian Toman), legal status (documented vs undocumented), length of stay (<5 years vs ≥5 years), parity, husband employment, and education. It also contained questions asking about the intimate partner violence (ever been physically, emotionally, or sexually abused during the recent pregnancy) poor mental health (feeling down, depressed, or hopeless), housing issues (housing issues such as unaffordability, lack of appropriate furniture, home insecurity in last year), food insecurity (missing a meal, while hungry, due to not enough food/no money to buy during last month), and the number of antenatal care (ANC) visits. The optimal number of ANC was ≥ 8 visits during pregnancy based on the WHO recommendation (2016) (18). The adverse pregnancy outcomes included preterm labor, abortion, stillbirth, eclampsia/pre-eclampsia, early rupture of membrane, gestational hypertension, gestational diabetes, intrapartum Hemorrhage, infections, or any of the above-complications.

### Data collection

A pilot study, including 12 participants was performed before the main study to assess the questionnaire comprehensibility and appropriateness and to ensure that the questions were well defined, clearly understood, and presented consistently. The interview session took place at a separate room in the community health center where only the interviewer and interviewee were present. The interviewers were three Afghan graduate midwifery female students at Tehran University of Medical Sciences. The participants were approached by the interviewers during the visit to the community health center. A brief introduction to the study and its objectives was provided for each participant before the interview. Given verbal consent, the woman was entered the study and written consent was obtained before the interview. Data were collected in the questionnaire through face-to-face interview in a separate room at the community health center in which only the interviewer and the participants were present. Interviews were conducted in the Persian language and each lasted approximately 15 minutes. At the end of the interview, the women’s concern regarding the adverse pregnancy outcomes and how to prevent them were addressed by the interviewers.

### Data analysis

The distribution of sociodemographic characteristics and the prevalence of adverse pregnancy outcomes, poor ANC, intimate partner violence, poor mental health, housing issues, and food insecurity were reported using descriptive statistics (Table 1 and 2). The association between sociodemographic factors, poor ANC, intimate partner violence, poor mental health, housing issues, and food insecurity with adverse pregnancy outcomes were assessed using the chi-square test and reporting the crude odds ratios (Tables 3 and 4). Subsequently, the adjusted odds ratios were calculated using logistic models to account for the potential confounding effects of sociodemographic factors (Tables 3 and 4). The STATA software version 14 was used for data analysis. P-value less than 0.05 was determined as the significant statistical level.

**Table 1.**
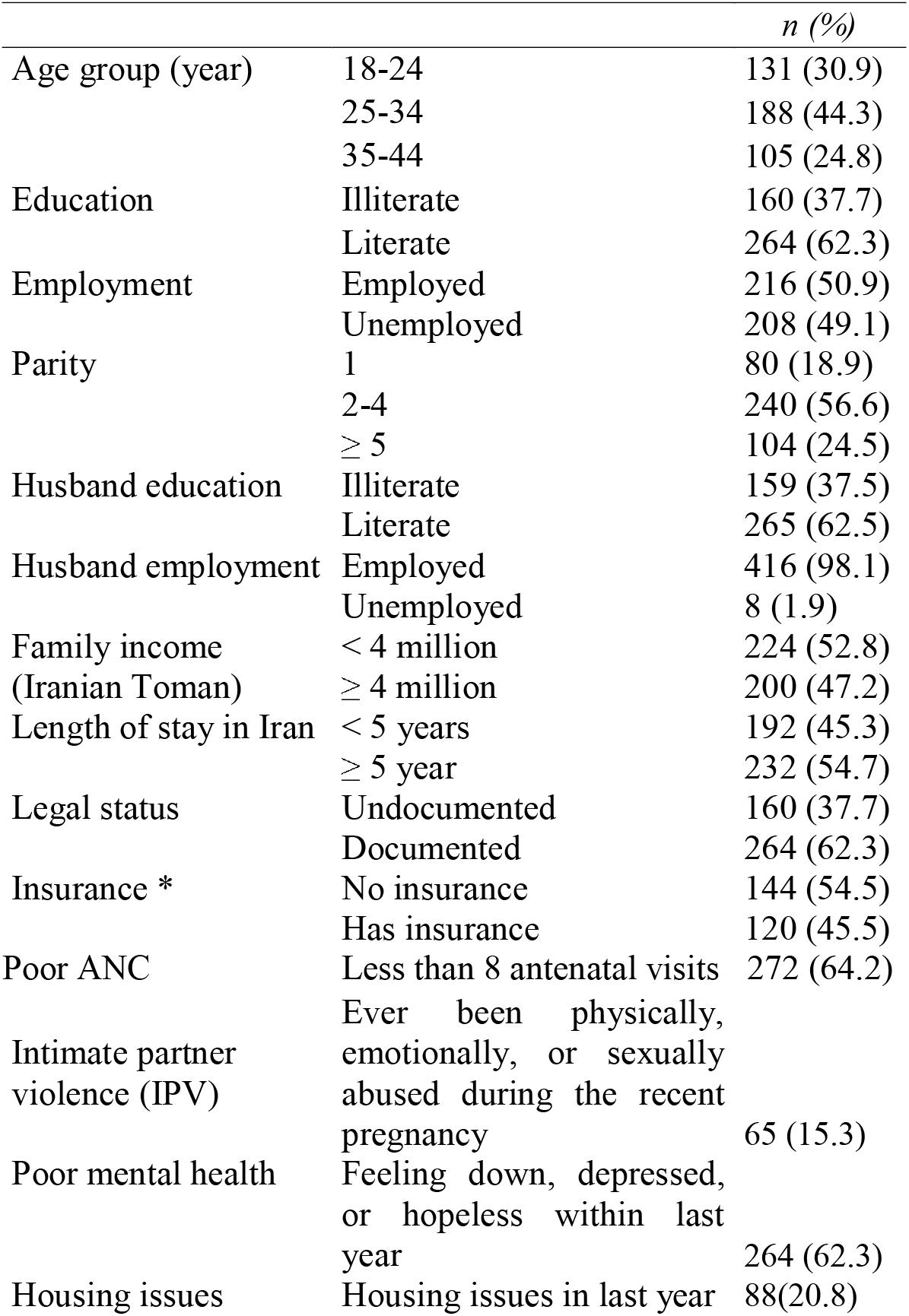
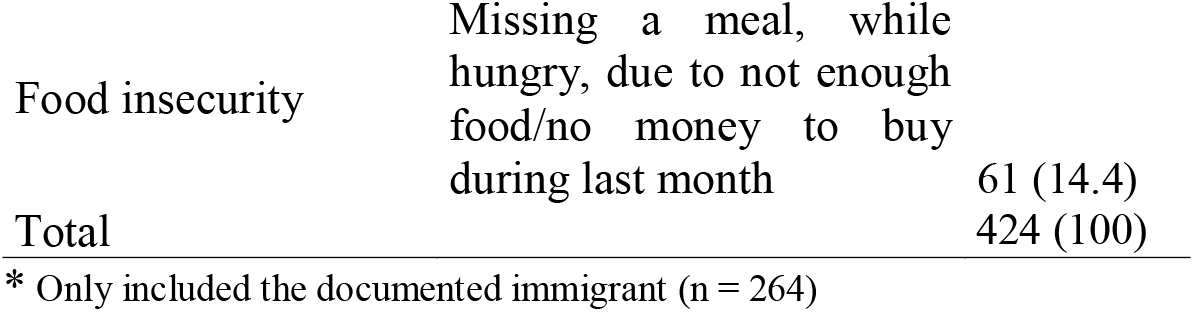
The distribution of the participant’s characteristics (n=424)

|  |  | *n (%)* |
| --- | --- | --- |
| Age group (year) | 18-24 | 131 (30.9) |
|  | 25-34 | 188 (44.3) |
|  | 35-44 | 105 (24.8) |
| Education | Illiterate | 160 (37.7) |
|  | Literate | 264 (62.3) |
| Employment | Employed | 216 (50.9) |
|  | Unemployed | 208 (49.1) |
| Parity | 1 | 80 (18.9) |
|  | 2-4 | 240 (56.6) |
|  | ≥ 5 | 104 (24.5) |
| Husband education | Illiterate | 159 (37.5) |
|  | Literate | 265 (62.5) |
| Husband employment | Employed | 416 (98.1) |
|  | Unemployed | 8 (1.9) |
| Family income (Iranian Toman) | < 4 million | 224 (52.8) |
|  | ≥ 4 million | 200 (47.2) |
| Length of stay in Iran | < 5 years | 192 (45.3) |
|  | ≥ 5 year | 232 (54.7) |
| Legal status | Undocumented | 160 (37.7) |
|  | Documented | 264 (62.3) |
| Insurance * | No insurance | 144 (54.5) |
|  | Has insurance | 120 (45.5) |
| Poor ANC | Less than 8 antenatal visits | 272 (64.2) |
| Intimate partner violence (IPV) | Ever been physically, emotionally, or sexually abused during the recent pregnancy | 65 (15.3) |
| Poor mental health | Feeling down, depressed, or hopeless within last year | 264 (62.3) |
| Housing issues | Housing issues in last year | 88(20.8) |

**Table 2.** The prevalence of the adverse outcome in the recent pregnancy in the study population (n=424)

|  | n (%) |
| --- | --- |
| Preterm Labor | 88 (20.8) |
| Abortion | 24 (5.7) |
| Stillbirth | 32 (7.5) |
| Eclampsia/pre-eclampsia | 16 (3.8) |
| Early rupture of membrane | 48 (11.3) |
| Gestational hypertension | 64 (15.1) |
| Gestational diabetes | 56 (13.2) |
| Intrapartum Hemorrhage | 54 (12.7) |
| Infections | 168 (39.6) |
| Any of above-complications | 250 (56.6) |

**Table 3.**
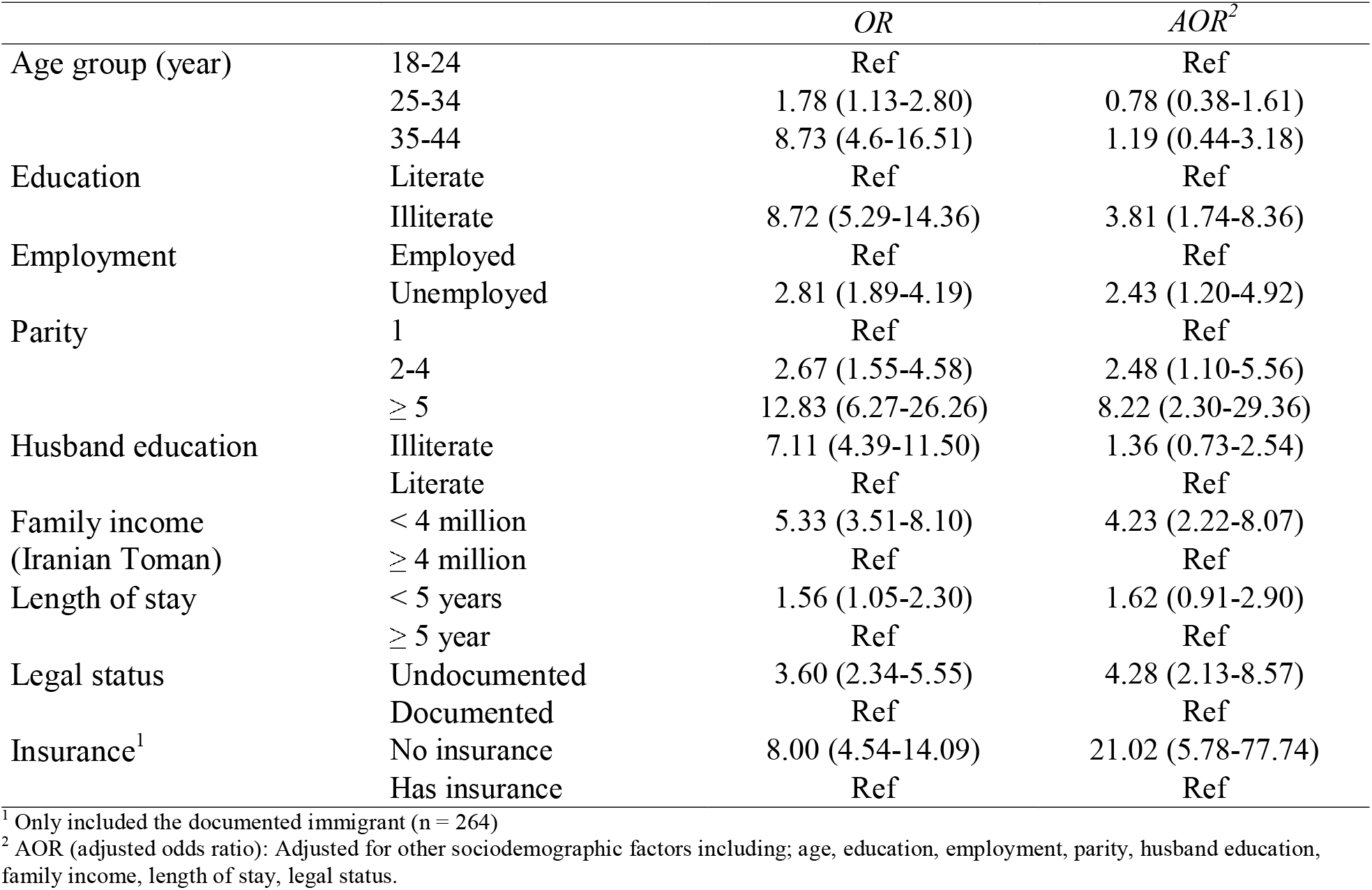
The association of sociodemographic characteristics with adverse pregnancy outcome among Afghan women (n=424)

|  |  | *OR* | *AOR*<sup>2</sup> |
| --- | --- | --- | --- |
| Age group (year) | 18-24 | Ref | Ref |
|  | 25-34 | 1.78 (1.13-2.80) | 0.78 (0.38-1.61) |
|  | 35-44 | 8.73 (4.6-16.51) | 1.19 (0.44-3.18) |
| Education | Literate | Ref | Ref |
|  | Illiterate | 8.72 (5.29-14.36) | 3.81 (1.74-8.36) |
| Employment | Employed | Ref | Ref |
|  | Unemployed | 2.81 (1.89-4.19) | 2.43 (1.20-4.92) |
| Parity | 1 | Ref | Ref |
|  | 2-4 | 2.67 (1.55-4.58) | 2.48 (1.10-5.56) |
|  | ≥ 5 | 12.83 (6.27-26.26) | 8.22 (2.30-29.36) |
| Husband education | Illiterate | 7.11 (4.39-11.50) | 1.36 (0.73-2.54) |
|  | Literate | Ref | Ref |
| Family income<br>(Iranian Toman) | < 4 million | 5.33 (3.51-8.10) | 4.23 (2.22-8.07) |
|  | ≥ 4 million | Ref | Ref |
| Length of stay | < 5 years | 1.56 (1.05-2.30) | 1.62 (0.91-2.90) |
|  | ≥ 5 year | Ref | Ref |
| Legal status | Undocumented | 3.60 (2.34-5.55) | 4.28 (2.13-8.57) |
|  | Documented | Ref | Ref |
| Insurance<sup>1</sup> | No insurance | 8.00 (4.54-14.09) | 21.02 (5.78-77.74) |
|  | Has insurance | Ref | Ref |

<sup>1</sup> Only included the documented immigrant (n = 264)
<sup>2</sup> AOR (adjusted odds ratio): Adjusted for other sociodemographic factors including: age, education, employment, parity, husband education, family income, length of stay, legal status.

**Table 4.** The impact of domestic violence, poor mental health, housing issues, and food security on pregnancy complications (n=424)

|  | Poor ANC | IPV | Poor mental health | Housing issues | Food security |
| --- | --- | --- | --- | --- | --- |
| Any pregnancy complications |  |  |  |  |  |
| OR (95% CI) | 12.19 (7.54-19.69) | 6.85 (3.18-14.76) | 9.36 (5.95-14.78) | 11.00 (5.16-23.47) | 6.24 (2.88-13.49) |
| Adjusted OR* | 10.50 (5.40-20.39) | 2.72 (1.10-6.77) | 4.77 (2.54-8.94) | 1.45 (0.48-4.42) | 3.35 (1.34-8.36) |

\* Adjusted for possible confounding factors including household income, legal status, mother's age, education, and employment, husband's education

### Ethics approval and consent to participate

The protocol of the present study was reviewed and approved by the Institutional Review Board (IRB) of both Graduate School of Medicine, Kyoto University (Ethic code: R1836), and Tehran University of Medical Science (Ethic code: 1397.945). The objectives of the study, potential harms, and benefits were explained for all participants and informed written consent was obtained thereafter.

## Results

Table 1 indicates the sociodemographic characteristics of Afghan women in the present study. A total of 424 women aged 18-44 years old enrolled in this study. All the participants who consent to attend the interview completed the study. Almost a third (37.7%) of them were illiterate and half of them were employed. The majority had between 2 to 4 pregnancy experiences. The household income was less than 4 million (Iranian Toman) in approximately half of the women (52.8%). Almost two-thirds (62.3%) of the women were documented immigrants. Amongst documented immigrants, less than half (45.5%) had health insurance. Approximately two-thirds (64.2%) of women reported having equal or more than 8 ANC visits during their recent pregnancy. Sixty-five women (15.3%) reported being physically, emotionally, or sexually abused during their recent pregnancy. More than 60% of Afghan women reported depression symptoms such as feeling down or hopeless during last year. Housing issues were reported by 88 (20.8%) women within last year. Food insecurity was documented among 61 (14.4%) of Afghan women enrolled in this study.

### Prevalence of pregnancy complications

The most prevalent obstetrics complication was preterm labor observed in 88 (20.8%) of women in this study. Gestational hypertension (15.1%), gestational diabetes (13.2), intrapartum hemorrhage (12.7%), early rupture of membrane (11.3%), stillbirth (7.5%), and abortion (5.7%) were the other reported obstetric complications. Gynecologic infection was reported by 168 (39.6%) of women. Overall, 250 (56.6%) of participants reported at least one pregnancy complication during their recent pregnancy.

### The association of sociodemographic factors with adverse pregnancy outcomes

Although in bivariate analysis, the higher age was associated with more pregnancy complications; adjusting for other variables, diluted this association (Table 3). The illiterate and unemployed women were more likely to experience adverse pregnancy outcomes (AOR = 3.81; 95% CI [1.74-8.36] and AOR = 2.43; 95% CI [1.20-4.92], respectively). The women with higher parity were more likely to experience adverse pregnancy outcome (Table 3). Husband illiteracy was associated with higher risk of adverse outcomes (OR = 7.11; 95% CI [4.39-11.5]); however, this association melted after adjustment for other sociodemographic factors (AOR = 1.36; 95% CI [0.73-2.54]). Household income appeared to be lower than 4 million/month in women who experienced any adverse pregnancy outcomes (AOR = 4.23; 95% CI [2.22-8.07]). Recent Afghan immigrants (equal or less than 5 years) reported higher pregnancy complications compared to settled ones (more than 5 years), although this association was not significant after multivariable adjustment (AOR = 1.62; 95% CI [0.91-2.90]). The undocumented immigrants were more likely to experience adverse pregnancy, even after adjustment for other variables (AOR = 4.28; 95% CI [2.13-8.57]). Among documented immigrants, those with no health insurance had higher risk of any adverse pregnancy outcomes, this association almost doubled after adjustment for other sociodemographic variables (OR = 8.00; 95% CI [4.54-14.09] versus AOR = 21.02; 95% CI [5.78-77.74]).

### The impact of IPV, poor mental health, housing issues, and food security on pregnancy outcome

As Table 4 indicates, the Afghan women who experienced IPV (physically, emotionally, and sexually) were more likely to experience adverse pregnancy outcomes in both bivariate and multivariate analysis (AOR = 2.72; 95% CI [1.10-6.77]). Poor mental health appeared to be associated with higher occurrence of pregnancy complications in Afghan women enrolled in this study (AOR = 4.77; 95% CI [2.54-8.94]). Although housing issues were more reported in women with higher adverse outcomes (AOR = 11.00; 95% CI [5.16-23.47]), this association melted after adjustment for sociodemographic factors (AOR = 1.45; 95% CI [0.48-4.42]). The unavailability of appropriate amounts of food for households appeared to be a significant factor influencing the occurrence of adverse pregnancy outcomes in Afghan women (AOR = 3.35; 95% CI [1.34-8.36]).

### The association between poor ANC and adverse pregnancy outcome

The inadequate ANC (<8 visits) was significantly associated with adverse pregnancy outcomes in both bivariate and multivariate analyses (OR = 12.19; 95% CI [7.54-19.69], AOR = 10.50; 95% CI [5.40-20.39].

## Discussion

Afghan refugee women are a very vulnerable population in Iran whose health needs have been neglected for decades. A number of reasons have been cited as the obstacles toward adequate health care among them such as financial struggles, health illiteracy, gender inequality, gender-based violence, and lack of autonomy (6, 8, 11). The limited access to health care contributes to the higher incidence of adverse health outcomes, particularly during pregnancy, and could jeopardize the health of the mother and new borne (18). In the present study, we estimated the prevalence of adverse pregnancy outcomes and explored the associated sociodemographic factors as well as the impact of poor ANC, poor mental health, IPV, food insecurity, and housing issues on pregnancy outcome in Afghan women living in Iran. More than half of the participants in the present study reported at least one adverse pregnancy outcome in their recent pregnancy. The prevalence of adverse pregnancy outcomes appeared to be higher in Afghan compared to Iranian women (almost two times higher for preterm labor, pre-eclampsia, gestational hypertension, and diabetes) (19-21). The results showed that undocumented, illiterate, and unemployed Afghan women with low socioeconomic status are more likely to experience adverse pregnancy outcomes. Furthermore, we observed lower prevalence of adverse pregnancy outcomes among documented immigrants with health insurance. It is also been found that food insecurity, poor ANC, IPV, and poor mental health could adversely impact the pregnancy outcome and we observed higher incidence of adverse outcomes among those suffering from these situations.

For many years, Iran has been a popular destination for a large number of Afghan refugees. The documents from UNHCR indicate that documented Afghan migrants have free access to primary health care similar to Iranian citizens (22) and benefit the free public health relevant programs such as vaccination and family-planning services. However, there are serious shortcomings in health provision for Afghan women such as lack of access to free ANC on contrary to Iranian women (23). Several studies, similar to our study, linked the inadequate ANC to adverse pregnancy outcomes. The lower socioeconomic, inadequate health literacy, and undocumented status could contribute to lower access to ANC and subsequently higher incidence of adverse pregnancy outcomes in immigrant and refugee women (24, 25).

Since 2016, all registered Afghan migrants could benefit the Universal Public Health Insurance in Iran. This substantially increased their access to the health system ever since (3); however, some vital ANC services such as prenatal screening tests or ultrasound are not covered and reimbursed by this insurance. This could reduce Afghan women’s access to adequate maternity care and increase the incidence of adverse pregnancy outcomes (6). Likewise, we found lower pregnancy complications among those documented migrants owning public health insurance, even after adjustment for other sociodemographic variables. On the other hand, the situation for undocumented migrants is unclear, we believe that the lower socioeconomic status and literacy along with lack of health insurance could considerably reduce the access of unregistered Afghan women to the health system and adequate antenatal care and lead to adverse pregnancy outcomes. Therefore, we advise the central government and international parties to provide affordable universal health coverage for all Afghan nationals, at least for Afghan women, regardless of their documented status to ensure the health of Afghan mothers and their offspring.

Food insecurity was associated with higher adverse pregnancy outcomes in the present study. A similar observation was documented among African immigrants in developed countries (26) and Latino immigrants in the USA (27). High prevalence of food insecurity (61%) has been observed in Afghan households in a study conducted in Tehran and Mashhad by Omidvar et al (15). Another study reported an even higher rate of food insecurity among the Afghan households in Pakdasht (88%) (28); however, in our study, the prevalence of food insecurity was much lower (14.4%). This could be due to the difference in study setting and measurement. Several factors contribute to the food insecurity in Afghan households such as low levels of literacy, illegal residential status, unemployment, housing issues, and low socioeconomic status (15). The lack of adequate calories, minerals, and vitamins in pregnant women could result in lower birth weight, preterm birth, premature rupture of membrane, and higher rate of infections (29). This calls for interventions and initiatives to improve food security in Afghan households and to provide adequate mineral and vitamin supplements for pregnant mothers.

Intimate partner violence is an inevitable widespread phenomenon among the refugee population (30). IPV was reported by almost 15% of Afghan women enrolled in our study. A similar rate was reported for Iranian women living in Mazandaran province in Iran (12). Low literacy and socioeconomic factors contribute to the higher IPV incidence against women (5, 12). We also found a significant association between IPV and adverse pregnancy outcomes. Although the knowledge regarding the IPV and associated factors among Afghan women living in Iran is limited, the existent literature indicates higher prevalence of IPV in Afghan compared to Iranian women (5). A study in Semnan province documented poor reproductive health in Afghan women suffering from IPV(5); however, our study is the first study that reports on the negative impact of IPV on adverse pregnancy outcomes in Afghan women living in Iran. The results emphasize the necessity of expanding the safety net to enhance the psychosocial support for Afghan women in Iran.

Poor mental health and psychosocial stress are independent risk factors of adverse pregnancy outcomes (16); however, the impact could be modified by culture, ethnic, and religious practices (31, 32). In our study, there was a significant association between poor mental health and adverse pregnancy outcomes. The feeling of discrimination, abandonment, and isolation along with employment and financial issues cause enormous psychosocial distress and could impair the physical and mental integrity and lead to mental illnesses such as anxiety and depression (33). Besides, the studies have shown increased vulnerability in female refugees and immigrants, particularly those at the young ages (34, 35). The negative impact of poor mental health could more evident in pregnant women who may already be overwhelmed with the psychological burden of maternity care (17). This highlights the importance of psychological counseling in maternity care.

### Limitation

Although this is the first study that reports the prevalence, associated factors, and the impact of food insecurity, poor mental health, and intimate partner violence on pregnancy outcomes in Afghan women living in Iran, there were some shortcomings should be considered in interpreting the results and implementing future studies. First, since the main objectives of this survey were to explore the barriers toward adequate ANC in Afghan women living in Iran (6, 8), we were not able to use standard questionnaires to collect the data on intimate partner violence, poor mental health, housing issues, and food security due to limited resources and time; instead, we included some key short questions asking about intimate partner violence “have you ever been physically, emotionally, or sexually abused during the recent pregnancy?”, poor mental health “feeling down, depressed, or hopeless within last year”, housing issues “housing issues in the last year”, and food insecurity “missing a meal, while hungry, due to not enough food/no money to buy during last month” to account for such shortcomings. Second, we sampled the women who visit the community health centers to receive ANC in an urban setting and the results cannot be extrapolated to the suburb and rural Afghan population. This could also be one of the reasons for lower rates of some indicators such as IPV or food insecurity in our study. Future studies should account for such discrepancies in sociodemographic factors between urban and rural populations.

## Conclusion

To our knowledge, this is the first study that explored the prevalence and associated factors of adverse pregnancy outcomes and the impact of intimate partner violence, food insecurity, poor mental health on pregnancy outcomes among Afghan women in Iran. The results showed higher prevalence of adverse pregnancy outcomes in Afghan compared to Iranian women. The low literacy and socioeconomic status, unemployment, illegal status, and lack of health insurance were some of the important indicators of adverse pregnancy outcomes in Afghan women. Food insecurity, poor mental health, IPV, and poor ANC were significantly associated with higher incidence of pregnancy complications in Afghan women. Enhancing the psychosocial support and empowering Afghan women through expanding the social network and safety net should be a priority for the central government and international parties. Psychological counseling should be incorporated into routine maternity care for Afghan refugees. Access to free antenatal care is a right for all Afghan women and it should be facilitated by universal health insurance for all Afghans regardless of their legal status.

## Data Availability

Data could be available upon a reasonable request and with the permission of Tehran University of Medical Science ethical committee.

## Competing interests

The authors declare no conflict of interests.

## Funding

There was no source of funding for the present study.

## Authors’ contributions

OD, TN, MK, and MOK contributed to the study conception and design, Data analysis and interpretation, and Critical revision of the article. OD, FD, SS, and conducted the interviews and collect the data. OD, TN, MK, and FD wrote and revised the first draft. All authors read and approved the final manuscript.

## Acknowledgment

We would like to thank our distinguished colleagues at Tehran University of Medical Science, whose precious efforts helped us to a great extent in implementing this study.

## Notes

### Competing Interest Statement

The authors have declared no competing interest.

### Funding Statement

The authors recieved no fund for present study.

### Author Declarations

The protocol of the present study was reviewed and approved by the Institutional Review Board (IRB) of both Graduate School of Medicine, Kyoto University (Ethic code: R1836), and Tehran University of Medical Science (Ethic code: 1397.945).

## References

1. UNHCR. Islamic Republic of Iran, Solutions Strategy for Afghan Refugees UNHCR. 2014a.

2. Abbasi-Shavazi M. Return to Afghanistan? A study of Afghans living in Mashhad, Islamic Republic of Iran. Afghanistan Research and Evaluation Unit; 2005.

3. Hosseini Divkolaye NS, Burkle FM, Jr. The Enduring Health Challenges of Afghan Immigrants and Refugees in Iran: A Systematic Review. PLoS Curr. 2017;9:ecurrents.dis.449b4c549951e359363a90a7f4cf8fc4.

4. IOM. International Migration Report. United Nations; 2018.

5. Delkhosh M, Merghati Khoei E, Ardalan A, Rahimi Foroushani A, Gharavi MB. Prevalence of intimate partner violence and reproductive health outcomes among Afghan refugee women in Iran. Health Care for Women International. 2019;40(2):213–37.

6. Dadras O, Taghizade Z, Dadras F, Alizade L, Seyedalinaghi S, Ono-Kihara M, et al. “It is good, but I can’t afford it …” potential barriers to adequate prenatal care among Afghan women in Iran: a qualitative study in South Tehran. BMC Pregnancy and Childbirth. 2020;20(1):274.

7. Otoukesh S, Mojtahedzadeh M, Sherzai D, Behazin A, Bazargan-Hejazi S, Bazargan M. A retrospective study of demographic parameters and major health referrals among Afghan refugees in Iran. Int J Equity Health. 2012;11:82.

8. Dadras O, Dadras F, Taghizade Z, Seyedalinaghi S, Ono-Kihara M, Kihara M, et al. Barriers and associated factors for adequate antenatal care among Afghan women in Iran; findings from a community-based survey. BMC Pregnancy and Childbirth. 2020;20(1):427.

9. Small R, Yelland J, Lumley J, Brown S, Liamputtong P. Immigrant Women’s Views About Care During Labor and Birth: An Australian Study of Vietnamese, Turkish, and Filipino Women. Birth. 2002;29(4):266–77.

10. Higginbottom GMA, Morgan M, Alexandre M, Chiu Y, Forgeron J, Kocay D, et al. Immigrant women’s experiences of maternity-care services in Canada: a systematic review using a narrative synthesis. Systematic Reviews. 2015;4(1):13.

11. Mohammadi S, Carlbom A, Taheripanah R, Essén B. Experiences of inequitable care among Afghan mothers surviving near-miss morbidity in Tehran, Iran: a qualitative interview study. International Journal for Equity in Health. 2017;16(1):121.

12. Abdollahi F, Abhari FR, Delavar MA, Charati JY. Physical violence against pregnant women by an intimate partner, and adverse pregnancy outcomes in Mazandaran Province, Iran. J Family Community Med. 2015;22(1):13–8.

13. Ivers LC, Cullen KA. Food insecurity: special considerations for women. The American Journal of Clinical Nutrition. 2011;94(6):1740S–4S.

14. Food Insecurity And Health Outcomes. Health Affairs. 2015;34(11):1830–9.

15. Omidvar N, Ghazi-Tabatabie M, Sadeghi R, Mohammadi F, Abbasi-Shavazi MJ. Food insecurity and its sociodemographic correlates among Afghan immigrants in Iran. J Health Popul Nutr. 2013;31(3):356–66.

16. Hobel C, Culhane J. Role of Psychosocial and Nutritional Stress on Poor Pregnancy Outcome. The Journal of Nutrition. 2003;133(5):1709S–17S.

17. Witt WP, Wisk LE, Cheng ER, Hampton JM, Hagen EW. Preconception Mental Health Predicts Pregnancy Complications and Adverse Birth Outcomes: A National Population-Based Study. Maternal and Child Health Journal. 2012;16(7):1525–41.

18. Laditka SB, Laditka JN, Mastanduno MP, Lauria MR, Foster TC. Potentially avoidable maternity complications: an indicator of access to prenatal and primary care during pregnancy. Women Health. 2005;41(3):1–26.

19. Tavakolipour S, Beigi M, Nekuei N, Shafiei F. The Prevalence of Pregnancy Hypertensive Disorders and Their Related Factors in the Second and Third Level Hospitals Affiliated to Isfahan University of Medical Sciences, Isfahan, Iran. Journal of Midwifery and Reproductive Health. 2019;7(3):1726–31.

20. Kharaghani R, Cheraghi Z, Okhovat Esfahani B, Mohammadian Z, Nooreldinc RS. Prevalence of Preeclampsia and Eclampsia in Iran. Arch Iran Med. 2016;19(1):64–71.

21. Sharifi N, Khazaeian S, Pakzad R, Fathnezhad Kazemi A, Chehreh H. Investigating the Prevalence of Preterm Birth in Iranian Population: A Systematic Review and Meta-Analysis. J Caring Sci. 2017;6(4):371–80.

22. IUNHCR. UNHCR Global Appeal 2011 Update: United Nations High Commissioner for Refugees (UNHCR); 2011b [Available from: http://www.unhcr.org/4cd96d099.html.

23. Tober DM, Taghdisi MH, Jalali M. “Fewer children, better life” or “as many as God wants”? Family planning among low-income Iranian and Afghan refugee families in Isfahan, Iran. Med Anthropol Q. 2006;20(1):50–71.

24. Reed MM, Westfall JM, Bublitz C, Battaglia C, Fickenscher A. Birth outcomes in Colorado’s undocumented immigrant population. BMC Public Health. 2005;5(1):100.

25. Korinek K, Smith KR. Prenatal care among immigrant and racial-ethnic minority women in a new immigrant destination: Exploring the impact of immigrant legal status. Social Science & Medicine. 2011;72(10):1695–703.

26. Lindsay KL, Gibney ER, McAuliffe FM. Maternal nutrition among women from Sub-Saharan Africa, with a focus on Nigeria, and potential implications for pregnancy outcomes among immigrant populations in developed countries. Journal of Human Nutrition and Dietetics. 2012;25(6):534–46.

27. Sano Y, Garasky S, Greder KA, Cook CC, Browder DE. Understanding Food Insecurity Among Latino Immigrant Families in Rural America. Journal of Family and Economic Issues. 2011;32(1):111-23.

28. Abdollahi M, Abdollahi Z, Sheikholeslam R, Kalantari N, Kavehi Z, Neyestani TR. High occurrence of food insecurity among urban Afghan refugees in Pakdasht, Iran 2008: a cross-sectional study. Ecol Food Nutr. 2015;54(3):187–99.

29. Laraia BA, Siega-Riz AM, Gundersen C. Household Food Insecurity Is Associated with Self-Reported Pregravid Weight Status, Gestational Weight Gain, and Pregnancy Complications. Journal of the American Dietetic Association. 2010;110(5):692–701.

30. Garcia-Moreno C, Jansen HA, Ellsberg M, Heise L, Watts CH. Prevalence of intimate partner violence: findings from the WHO multi-country study on women’s health and domestic violence. Lancet. 2006;368(9543):1260–9.

31. Thomas PE, Beckmann M, Gibbons K. The effect of cultural and linguistic diversity on pregnancy outcome. Australian and New Zealand Journal of Obstetrics and Gynaecology. 2010;50(5):419–22.

32. Hollander A-C, Bruce D, Burström B, Ekblad S. The Association Between Immigrant Subgroup and Poor Mental Health: A Population-Based Register Study. The Journal of Nervous and Mental Disease. 2013;201(8).

33. Beiser M, Hou F. Mental Health Effects of Premigration Trauma and Postmigration Discrimination on Refugee Youth in Canada. The Journal of Nervous and Mental Disease. 2016;204(6).

34. Pernice R, Brook J. Refugees’ and Immigrants’ Mental Health: Association of Demographic and Post-Immigration Factors. The Journal of Social Psychology. 1996;136(4):511–9.

35. Ellis B, MacDonald H, Klunk-Gillis J, Lincoln A, Strunin L, Cabral H. Discrimination and mental health among Somali refugee adolescents: The role of acculturation and gender. American Journal of Orthopsychiatry. 2010;84 (4):564–75.

